# ‘Journeys’ to mental health and HIV wellness: adapting a community-based group program from Nigeria to Washington, DC

**DOI:** 10.64898/2026.07.29.26359227

**Authors:** Julie Pulerwitz, Ann Gottert, Adedotun Ogunbajo, Arona Dieng, Jennifer Kwait, Cody Henry, Chrisitan Morris, DeMarc Hickson

## Abstract

**Background:** Evidence-based programs are needed to address the multiple stigmas and disproportionately poor mental health and HIV-related outcomes that marginalized populations often face. In this implementation science study, we adapted a community-based program that we originally developed and tested in Lagos, Nigeria, to the Washington, DC context.

**Methods:** We used the ADAPT-ITT framework to guide a multi-stage adaptation process in 2024. We conducted four serial focus group discussions with the same six Black gay or bisexual men (GBM), to review the original 4-session curriculum. We also conducted in-depth interviews with additional GBM (n=7), and other stakeholders (n=8) including local health department staff and HIV and mental health service providers, and held informal discussions with cognitive behavioral therapy (CBT) and stigma experts. We explored curriculum relevance, acceptability, feasibility, delivery format, and anticipated effects. Subsequent baseline surveys from a pilot evaluation with Black GBM (n=63) assessed psychosocial well-being using validated scales.

**Results:** Community members and other stakeholders considered the program acceptable, feasible, appropriate, and needed in the local context. Core elements from the original intervention were retained, including its HIV status-neutral approach (i.e., with activities appropriate both for HIV prevention and treatment), four-session structure, and delivery by community health educators within community-based organizations. Adapted content emphasized critical reflection around multiple stigmas and their internalization, affirming identities, CBT skill-building, and HIV self-care and wellness. Components promoting shared resilience were strengthened, inspiring the program name, *Journeys of Shared Resilience*. Baseline survey data highlighted high levels of depression and internalized stigma alongside high personal resilience, underscoring the salience of the program.

**Conclusions:** Using a systematic, community-partnered approach, we successfully adapted an evidence-based program for Black GBM in Washington, DC. *Journeys* represents a promising status-neutral approach to addressing multiple stigmas, mental health, and HIV prevention and care. Pilot testing is ongoing.

## INTRODUCTION

Stigmatizing attitudes and related discrimination are major drivers of various adverse mental and physical health outcomes, including depression, anxiety, and both HIV acquisition and low treatment use (1–4). Minority populations often face multiple stigmas simultaneously, such as those related to HIV status, sexual orientation, and race (e.g., 5, 6). These views can be *internalized* (also called self-stigma), and evidence shows that the internalization of stigma is strongly and consistently associated with negative outcomes(7). Internalized stigma inhibits the uptake of HIV services like pre-exposure prophylaxis (PrEP) for HIV prevention as well as antiretroviral therapy (ART) for HIV treatment(4, 8, 9).

There is a need for more evidence around programming that can address internalized stigma – including for minority populations that commonly experience disproportionately poor HIV and mental health outcomes. Developing and testing innovative interventions are crucial for reducing stigma’s negative effects and providing opportunities to enhance positive outcomes like social support and shared resiliency, or, shared positive responses or adaptations in the face of adversity(4, 10–12). Recent publications, for example, have called for adding to responses that support individuals in the psychological work of reducing internalizing stigma to also incorporate a broader approach into programming that promotes understanding of the social structures that perpetuate stigma(9).

The Washington, DC metro area has one of the highest HIV prevalence levels of any U.S. jurisdiction (with almost 2% living with HIV as of 2023), and HIV incidence was estimated at 24.5 new infections per 100,000 population during 2022–2023, more than twice the U.S. national incidence rate(13). Both gay and bisexual men (GBM), and Black adults, experience particularly high HIV prevalence and incidence in the DC area(14, 15). About two-thirds (67%) of GBM living with HIV are virally suppressed(14), and it is estimated that less than half of DC residents that would benefit from HIV prevention through pre-exposure prophylaxis (PrEP) access it(15). A 2022 DC needs assessment identified combating HIV stigma as a priority for local programming(12). The report identified providing trauma-informed care and creating ‘safe spaces’ for programming as among the most critical activities to implement(15).

A promising programmatic strategy uses cognitive behavioral therapy (CBT) in a group setting to reduce multiple self-stigmas and promote HIV wellness and resiliency. CBT is a structured, time-limited, and skills-based psychotherapy that helps individuals identify and modify unhelpful patterns of thinking and behavior to improve mental health outcomes. Globally, CBT is among the most impactful approaches for supporting mental health and reducing psychosocial distress(16–18). It has become common for CBT to be implemented in a group setting(19–21), which can provide opportunities for a safe space to discuss challenges and enhance connection to and support from peers. A group model may also have other advantages related to cost and institutional capacity, and hence scalability and sustainability, in that multiple clients are reached at the same time.

Experience with a group CBT program for GBM in Nigeria laid important groundwork for adapting it to Washington, DC(22). This program integrated content encouraging critical reflection around multiple stigmas, built cognitive behavioral therapy skills and resiliency in the face of stress and trauma, and encouraged social support as well as self-care related to mental health and HIV prevention/treatment. It entailed four (4) weekly sessions (each 2.5 hours in length, with 10-15 participants), facilitated by trained community health workers. As an HIV ‘status-neutral’ intervention, HIV status was not a factor in participant recruitment and status disclosure was optional during programming. The Nigeria-based program was itself informed by evidence-based group CBT programs tailored for GBM and / or for people living with HIV in Canada, China, and the United States(19–21).

A randomized controlled trial (RCT) assessing the Nigeria-based program was conducted in Lagos in 2022(22). The RCT included a delayed intervention group methodology with a total of 240 participants in the combined immediate and delayed groups, plus a 3-month follow-up for the immediate program group only. The evaluation showed feasibility, acceptability, and preliminary efficacy of the group-based CBT model(22). The great majority of participants (88%) attended all four sessions and expressed high program satisfaction (98%). This was an environment characterized by high levels of stigma experienced by GBM(23, 24), and qualitative findings with participants in both groups post intervention indicated enhanced self-confidence and resilience when facing stigma. Survey data demonstrated sustained and significant positive changes in internalized stigma, depression, and anxiety at the three-month follow-up.

The current paper describes an implementation science study to adapt the Nigeria-based program - now called *Journeys of Shared Resilience* - to the Washington, DC context. To guide the adaptation process we used the ADAPT-ITT *(Assessment, Decision, Adaptation, Production, Topical Experts, Integration, Training*, and *Testing)* framework (25). This paper describes the ADAPT portion of the framework. We also share findings from the subsequent pilot program’s baseline survey to provide insights into levels of psychosocial well-being among the participants. A subsequent paper will summarize the ITT portion, including the pilot evaluation results.

## MATERIALS AND METHODS

The team guiding the adaptation process consisted of both program staff and researchers. Specifically, it included senior staff from a CBO that provides mental health and HIV-related services and an HIV research / policy institute affiliated with a federally qualified health center (FQHC) - both serving large client populations in DC - as well as researchers with global implementation science, stigma, and HIV expertise who developed and evaluated the group program in Nigeria. Ethical approval for this study was obtained by the Population Council Institutional Review Board (protocol #1023), and all study participants provided written consent.

Adapting the program entailed conducting formative research with community members and other stakeholders such as service providers. The main objective was to assess program acceptability, appropriateness, and feasibility within the Washington, DC context, via applying the ADAPT-ITT framework.

The framework is a systematic, iterative methodology for tailoring evidence-based interventions to new cultural and contextual settings or populations. ADAPT comprises several distinct phases. These include an <u>Assessment</u> of the program via formative research to identify potential fit and recommended revisions, <u>Deciding</u> whether the program’s core elements are appropriate for that new context, <u>Adapting</u> the program accordingly based on the results of the assessment, and <u>Producing</u> a final version of the adapted program. During this process, the program is also informed by consultations with content experts (described in the framework as ‘<u>Topical’ experts)</u>.

The current adaptation process began with the study team reviewing the original program structure and curriculum. The team revised certain elements to better align with the local context, such as removing discussion of Nigeria and adding content about stigma related to race, which is more pertinent in the Washington, DC context. The review resulted in a draft discussed during the formative research phase described below. We also identified additional issues to explore further during the formative research, such as screening around mental health, and appropriate exercises to promote shared resilience.

### Procedures for the formative research

Eligible community member participants were 18 years or older, lived or worked in the DC metro area, and self-identified as a Black GBM. Participants were purposively recruited through existing client networks (a mix of people with and without HIV), as well as via community events and social media outreach. Recruitment flyers were posted online and at community venues.

We conducted a series of four (4) sequential FGDs with the same group of people (n = 6) held over two weeks. The main purpose of the FGDs was to review each of the four program sessions in detail and to discuss reactions to each of the exercises. Each FGD lasted about two hours, was held in-person in a private location, and was guided by two experienced facilitators who were also members of the study team. FGD participants received $40 per session to compensate them for travel and their time.

This was followed by IDIs (n = 7) with a different group of Black GBM to further explore program acceptability and themes that emerged during the FGDs. The IDIs were 45-75 minutes in length and conducted by a trained study team member. They were either held in-person in a private location or via Zoom, per the participants’ preference. IDIs were audio-recorded and subsequently transcribed. IDI participants also received $40.

We also conducted IDIs with other individuals perceived to have important insights into potential program adaptation, who were then recruited by study team members. These stakeholders included representatives from the DC Department of Health (DC-Health) and local HIV and mental health service providers (n = 8). These IDIs focused on program need and fit in the context of other existing services in the DC area. IDIs were conducted by a trained interviewer (also a study team member) via Zoom and lasted 30-45 minutes. They were audio-recorded and subsequently transcribed. These stakeholders did not receive compensation for participation.

The FGD and IDI guides addressed perceived acceptability, feasibility, and appropriateness of the program, among other topics. Questions examining program acceptability explored how the intervention’s objectives, structure, and content would resonate with potential program participants. Questions examining program feasibility assessed logistical and structural considerations, including recruitment strategies, participant retention, and program delivery mechanisms. Questions examining program appropriateness explored how well the program aligned with the cultural and contextual realities of the proposed audience.

Recruitment for the FGDs/IDIs described above began on February 1, 2024 and ended on September 14, 2024.

### Qualitative data analysis

A summary of each FGD and IDI was prepared by the facilitator or interviewer shortly after its completion. The study team reviewed these summaries to inform the ongoing edits to the program curriculum. Transcripts of audio recordings were also coded in qualitative data analysis software (Dedoose v9) by two study team members, using both topical codes (i.e., per interview guide questions) and emergent codes (i.e., relevant new ideas or topics that were not included in the guide). The study team then identified themes to further inform and guide program adaptation.

### Consulting with topical experts

Study team members held discussions with several stigma, HIV and CBT content experts (n=3). We also facilitated a two-hour discussion with ∼20 participants at the 2024 NAESM conference, an annual meeting that addressed the HIV-related needs of the Black community.

Each consultation solicited general reactions to an outline of the program structure and curriculum content and then focused on questions most relevant to the particular individual (e.g., best practices for group-based CBT, stigma measurement). These discussions were summarized via written notes.

### Baseline survey data on psychosocial well-being

After the curriculum was finalized, a program evaluation with a group of Black GBM began. The evaluation baseline survey (n=63) assessed levels of internalized stigma, depression, anxiety, resilience, and community connectedness, amongst other topics, and results are shared here to provide community context. Recruitment began on November 1, 2024 and ended on April 1, 2025. Participants completed the baseline survey online and received $25. Former formative research participants were not eligible for the program evaluation, and thus results from the baseline survey draw from a different community sample than the formative research.

Psycho-social variables in the baseline survey were measured utilizing previously validated scales, with alphas reported below from the current study sample. A 10-item version of the Connor-Davidson Resilience Scale (26) was used to measure psychological resilience (alpha: 0.90). Coping was assessed using the 4-item Brief Resilient Coping Scale (alpha: 0.79) (27). Community connectedness was assessed using Frost et al.’s 8-item scale (28) (alpha: 0.88).

Internalized stigma was assessed with sets of items related to three separate topics: sexual identity (6 items, alpha: 0.87), race (8 items, alpha: 0.89), and HIV status (5 items, alpha: 0.87). These items were adapted from Kalichman et al.’s Internalized AIDS-related Stigma scale(29) and a separate stigma scale focused on race (30).

Depression was assessed using the 8-item Patient Health Questionnaire (PHQ-8) scale (31)(alpha: 0.93). Anxiety was assessed with the 7-item General Anxiety Disorder (GAD-7) scale(32) (alpha: 0.95).

## RESULTS

### I. Assessment Phase

Qualitative findings are organized according to the core implementation outcomes of acceptability, feasibility, and appropriateness. We have integrated insights from topical experts into FGD and IDI results throughout.

#### Acceptability

The program was viewed as acceptable by both community members and other stakeholders. Perceived acceptability stemmed from the recognition of prevalent needs among Black GBM in the Washington, DC area to address mental health issues and self-stigmas and to bolster community connectedness.

Community members expressed how they regularly and historically experienced stigma, and that the program would fill a significant gap in the availability of safe spaces for Black GBM to process their experiences together. As one FGD participant remarked:

> *This program will get people to open up about internalized stigma, which is important to address. And having people share their vulnerability and have support in a group setting is great. (Community member)*

Other stakeholders echoed this positive reception of the program:

> *People are looking for a community, looking for assistance, but they might not know the best routes to ask for it or how to obtain it. Putting people in spaces who share various experiences can assist…I think [the proposed program is] a great method. (HIV service provider)*
>
> *Stigma is a thing…[and] that’s what you’re addressing. I think it’s also relevant when I see the piece around healthy social networks…how do I build healthy communities? How do I build a healthy self? I think these are all legitimate and timely issues. So, in my opinion, I think it’s needed. (HIV service provider)*

Respondents also shared their view that the program would likely increase HIV service use, such as the following opinion:

> *The conversation will get HIV wellness on people’s minds. And then they would hopefully feel empowered to make the next step in engaging in care, either for treatment or prevention. (HIV service provider)*

#### Group-based nature of the program

Participants consistently emphasized the advantages of mutual learning and support in a group setting. The topical expert on CBT approaches agreed, affirming that the content appears suitable – and not overly sensitive – for group discussion. When asked about the possibility of implementing groups that include people with a range of characteristics, most participants thought the benefits of diverse views and experiences outweighed any potential drawbacks. As a service provider stated:

> *let’s say, if you had somebody from the south versus the north…I don’t know your struggles. You don’t know my struggles. So yeah, I think it’s [a group program] a great way to actually be empathetic to other people. (mental health service provider)*

Another respondent similarly reflected:

> *A lot of the power in such a program like this is that you’re bringing together people from across the aisle. So I wouldn’t necessarily want to sit in a room [with] other people that experience life the same way that I do…having differing views is what’s really nice about this. (Community member)*

A few respondents suggested the idea of separating groups by age, citing different life experiences and perspectives. As one stated:

> *I would be very interested in seeing if there was a senior cohort and what that would look like, especially talking about internalized stigma. I mean, our seniors are the ones who lived through the HIV/AIDS crisis. (Government official)*

Privacy and confidentiality emerged as an area of concern related to the group-based program format, particularly due to the potential sensitivity of topics discussed. Both community members and other stakeholders emphasized the need for clear ground rules to protect participant disclosures and build trust. These measures were seen as essential for maintaining a safe and supportive environment for participants.

#### Inclusive HIV status neutral approach

Participants expressed a strong preference for integrating individuals regardless of HIV status within the same intervention groups, sometimes called a ‘status-neutral’ approach. It was seen as a strategy that fostered inclusivity and reduced stigma. A local government official described their rationale for supporting a status neutral approach:

> *It’ll help reduce the stigma around it because if you had like, this is the HIV group and this is the not HIV group, I think that’s stigmatizing….you’ll have some of the same issues in people who have HIV and people who don’t. (Government official)*

As one participant who is living with HIV also stated:

> *It’d be good to hear people’s different perspectives, like the HIV-negative people - they may have stigma about HIV that may not be true. And then somebody like me who’s living with it and born with it can educate them. (Community member)*

#### Feasibility

Most community members and stakeholders expressed confidence in the program’s feasibility, with many affirming that the design and delivery approach were likely to work well in the Washington, DC context.

#### Number and timing of sessions

We solicited feedback on the format used in Nigeria - four sessions lasting about 2.5 hours and implemented weekly. This was seen as feasible. Several participants noted the importance of spacing sessions out, to allow for reflection and practice of information and skills and bonding between participants. Participants suggested scheduling sessions on weekday evenings or weekends to accommodate participants’ availability.

One government stakeholder questioned whether a four-session program would provide sufficient time to fully address HIV treatment and prevention. This stakeholder proposed adding a session in a separate, safe space to discuss HIV-status-specific challenges among those living with HIV.

#### Engagement in sessions

Both providers and government stakeholders emphasized the importance of using strategies to support participant attendance, based on previous experience with implementing similar programs in DC. Recommended strategies included offering transportation support, such as gas or metro cards, and providing meals during sessions.

One service provider questioned whether a four-session program would provide sufficient time for participants to build trust and share personal experiences. Another government stakeholder suggested prioritizing trust-building activities among participants early in the program to create a foundation for open dialogue.

#### Facilitators of the sessions

The interviews explored opinions about who should facilitate these sessions. We probed about the pros and cons of having CBO staff who are members of the community, but not clinicians, as facilitators. The program in Nigeria included lay / community health workers (CHWs) as the main facilitators, with a trained psychologist available for additional support as needed. Most study participants responded positively to the idea of community-based facilitators. In addition, the topical expert in CBT shared the opinion that community-based program staff could successfully facilitate these types of sessions. As was stated:

> *I think community health workers know how to reach folks without using medical jargon that often gets confusing and are more relatable in this type of setting (HIV service provider)*
>
> *I don’t want…where someone feels like they can’t come because, “Oh, this person is going to give me a lecture.” (Community member)*

Further, many participants indicated a preference for program facilitators who shared lived experiences as Black GBM. The choice of facilitators was highlighted as a critical factor in ensuring the program’s cultural appropriateness. For example:

> *[The facilitator should be] a peer educator or community health worker…someone from the community. (FGD participant)*
>
> *Definitely somebody who is well connected to community…a facilitator who is empathetic and compassionate and can facilitate without spreading their own trauma into the midst. (Government health official)*

A few service providers and health department representatives - although not community member interviewees - recommended a combination of both mental health professionals and community member facilitators.

#### Safeguarding mental health of participants

The interviews also explored how to ensure that clinical supports were available when specific mental health needs or concerns of program participants arose during the sessions. Interviewees agreed with the approach of ensuring timely referrals to individual counseling or mental health services as needed.

The CBT expert also proposed considering screening out individuals struggling with serious mental health conditions from the group sessions. A different mental health service provider brought up the alternative of self-screening. An example of this could be, the provider stated, advising potential participants during screening/enrollment that the program was not intended or recommended for treatment of serious mental health conditions.

#### Appropriateness

The was general agreement by community members and other stakeholders that the program was appropriate for its intended population. Respondents noted program’s alignment with the cultural and social realities of Black GBM in Washington, DC.

Participants also provided suggestions for refinement of the curriculum, such as by addressing intersectional stigmas tied to race. This would be in addition to stigmas related to sexual orientation and HIV, which were emphasized in the Nigeria-based program. Recommendations also included incorporating local terminology and scenarios. One IDI participant related the following:

> *Cultural sensitivity [will be key]…as well as mental health awareness with regards to violence and/or traumatic experiences. So, using my own experience, for example, as a gay Black man living with HIV, who has also been affected with PTSD from gun violence since last year. (Community member)*

A new topic that was consistently identified as important to address in the local context was intra-community or intra-minority stigma – that is, stigma enacted and experienced within sub-communities of Black GBM. Participants felt that intra-community stigma was prevalent, particularly related to physical appearance, age, HIV status, and socio-economic status, and that it often translated into internalized stigma.

#### Continuity of the program after sessions end

Promoting connectedness and continuity among participants after program completion was seen as important by FGD and IDI respondents and was also emphasized by the topical expert group at the NAESM conference. Respondents felt it would be appropriate to invite participants in each group to exchange contact information and continue to connect after the sessions, as long as this was optional. The possibility of adding optional follow-up sessions to enhance program sustainability was also discussed.

#### Framing the program around building shared resilience

The CBT expert indicated that the promotion of shared resilience was a substantively unique aspect of this group-based program; that this topic was not typically a focus in group-based CBT approaches. Many participants agreed that the program component related to building shared resilience was a key strength. As a community member explained:

> *I’d definitely be able to learn from others’ experiences, but also give mine. And I think that would be beneficial for the program…being able to share on what we’ve done to be resilient…a lot of us will be grateful for that. (Community member)*

Community and other stakeholders described their definitions of shared resilience and how it could be promoted. Participants indicated how including the theme of shared resilience could strengthen the effects of the program both for program participants and more broadly. For example:

> *I love the concept of shared resiliency. It is better when people are aware others go through similar situations and can process together. (Mental health service provider)*
>
> *Once the program group realizes that they have shared experiences they start to bond – start working together to address issues. (HIV service provider)*
>
> *I feel like people want to be a part of something bigger and you’re creating more tools and smaller coalition blocks to build a larger coalition to make movement happen. (Government official)*

Suggestions for enhancing program activities to build shared resilience included incorporating arts-based activities, storytelling exercises to help participants process their experiences, and developing recommendations as a group for improvements in HIV, mental health and stigma reduction services in the local area.

### II. Decision Phase

Based on the results of the assessment phase, the study team concluded that the proposed strategy was acceptable, feasible, and appropriate for the local context. We confirmed that we would retain the original program’s core structure and would make several updates for the Washington, DC context (as described below.)

Thus, the program to be piloted would consist of four group sessions of about 2.5 hours each; group sizes of under 15-20 people each and ideally 10–12 people; sessions spaced over days or weeks; a status-neutral approach to HIV that did not restrict by HIV status and aimed to include both people living with and not living with HIV; groups that included people who represented a range of socio-demographic characteristics; and facilitation by trained community-based program staff with ready mental health referral pathways.

### III. Adaptation Phase

After reviewing the full formative research findings, the curriculum was further tailored. These adaptations were intended to ensure that the intervention remained both evidence-based and culturally responsive, supporting mental health, addressing internalized and external stigma, and fostering both HIV prevention and care engagement in the unique social context of Black GBM in Washington, DC.

Specific revisions to the curriculum were made to:

- Directly address intra-community and intra-minority stigma within examinations of stigma
- Incorporate visual and musical arts-based activities
- Include more programming that focused on uplifting and celebrating all aspects of identity
- Add new shared resilience-building exercises, emphasizing group storytelling and agency
- Remove a standalone activity focused on post-traumatic stress disorder, while integrating trauma awareness throughout other sessions
- Describe the HIV prevention and treatment program content as self-care, and position this component within the broader context of individual and group coping and resilience

### IV. Production Phase

**Figure 1** includes a summary of the structure and curriculum outline final program that was produced - called *Journeys of Shared Resilience.* A logo was also created with the aim of evoking the image of a collaborative journey.

**Figure 1.**
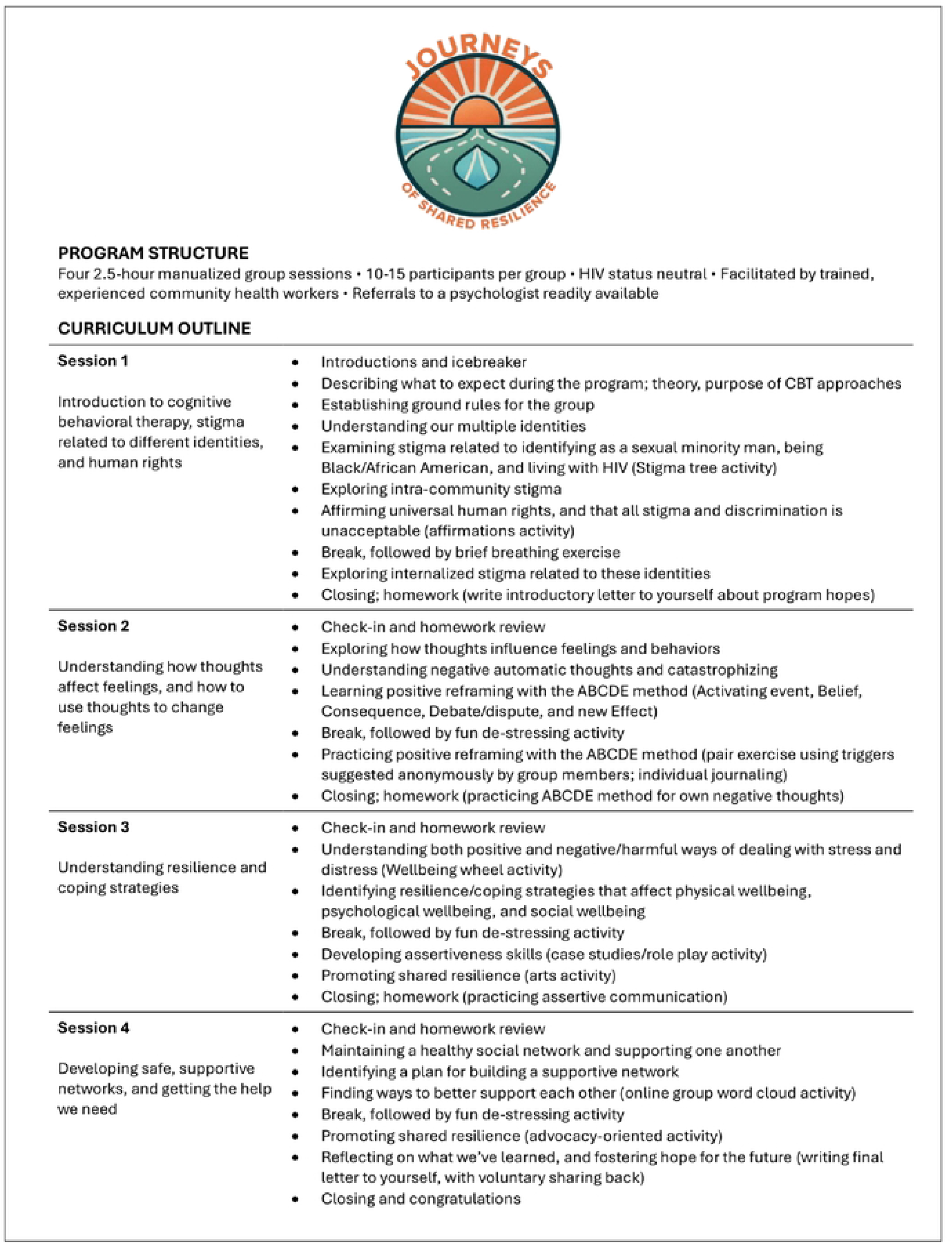
Journeys of Shared Resilience program summary.

### V. Psychosocial wellbeing from pilot baseline survey

In total, 63 Black GBM living the DC area participated in the baseline survey of the program evaluation, which took place from November 2024 until April 2025. Mean age was 38.4 years (range 20-79), and 31 participants (46%) reported living with HIV. **Figure 2** shows bar charts or histograms for four continuous variables related to psychosocial wellbeing. About half of the sample (44%) met the clinical cut-off for identifying likely cases of moderate or severe depression. About one quarter of the sample (26%) met the clinical cut-off for moderate or severe anxiety. Levels of reported internalized stigma were substantial, with the highest levels related to HIV status. Participants reported a wide range of responses regarding perceptions of coping, individual resilience, and community connectedness, with the highest levels found related to community connectedness. Responses suggest potential for improvement after program exposure.

**Figure 2.**
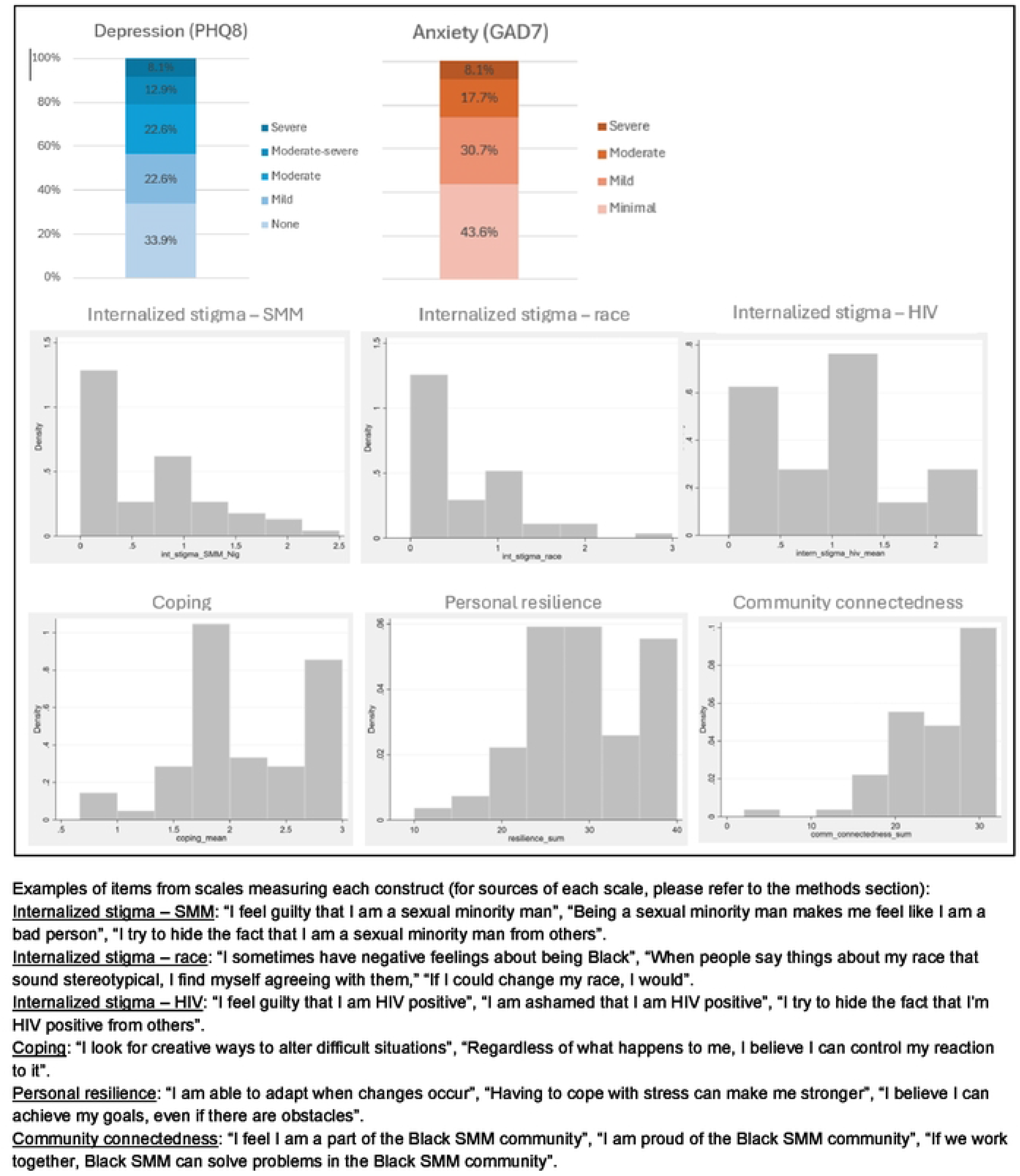
Baseline survey findings related to psychosocial wellbeing.

## DISCUSSION

This implementation science study details a systematic yet flexible adaptation of a group-based program—Journeys of Shared Resilience—to support mental health, address internalized stigma, and promote HIV prevention/care among Black GBM in Washington, DC. Our formative work demonstrates pathways for tailoring an evidence-based intervention developed for marginalized communities in Nigeria to communities in the US and highlights implications for future programming and research.

### Lessons from the Adaptation Process

Our academic-community partnership approach was critical in carrying out this process(33–35). Our multi-stage adaptation process, rooted in the ADAPT-ITT framework(25, 36), provided several opportunities for program refinement. Incorporating qualitative inputs from Black GBM enabled the integration of community needs and cultural nuances, particularly around the lived experiences of stigma, trauma, and resilience. Sequential FGDs with the same group of participants – combined with IDIs - allowed for a lens on the program as a whole, as well as individual perspectives. Additional input from local service providers and international experts was key for ensuring fidelity to core program components like CBT. All inputs maximized the likelihood of an acceptable, feasible, and appropriate program.

A recent systematic review of 37 health interventions for GBM concluded that programs typically did not sufficiently address the multiple, intersecting sources of stigma faced by the community, suggesting the need for this type of program(10). Another systematic review of 192 anti-stigma interventions found that few (∼5%) included marginalized populations such as sexual minorities, and few (∼5%) addressed stigma related to sexuality(37). Pilot baseline participants demonstrated a range of existing coping skills and indications of community connectedness—underscoring the potential to build on these strengths. However, consistent with broader literature on stigma as a driver of poor mental health, HIV acquisition, and care outcomes(1–4, 7, 8, 38), pilot participants revealed prevalent depression, anxiety, and internalized stigma, corroborating a need for psycho-social interventions in this setting.

By retaining original program elements such as the HIV status-neutral format, group-based delivery, and community-based facilitation, plus including explicit discussions of both shared resilience and intra-community stigma, the adapted curriculum responds directly to the needs and realities of Black GBM in DC.

### Implementation Strategy: CBT combined with shared resilience

The Journeys of Shared Resilience (‘Journeys’) program reinforces and extends the promise of group CBT as a strategy. There is growing evidence that group CBT is acceptable and appropriate for marginalized populations, such as sexual minorities(19, 22). What sets the current program apart is the explicit discussion of internalized and intersecting stigmas in a group setting, as well as the deliberate fusion of CBT strategies with community narrative sharing. The intervention both draws on and enhances personal and shared strengths, using a group space to model diverse coping mechanisms.

Notably, our adaptation process introduced new strategies to operationalize “shared resilience.” These included fostering vicarious coping (learning and practicing strategies seen in fellow participants) and celebrating individual identities. Such approaches accentuate wellness and collective agency, rather than exclusively focusing on distress. Feedback indicated that participants valued not only the individual mental health skills acquired but also the opportunities for social connection.

The program’s group model was delivered by community-based program staff with similarities in lived experience as participants. This is consistent with evidence suggesting peer and community-led facilitation can increase relatability, engagement, and scalability(39–41). As with our related study in Nigeria, acceptability and feasibility in Washington, DC were strongly linked to the trusted, relatable role of the facilitators.

### HIV Status Neutrality and Implications

An important feature of the Journeys program is its HIV status-neutral structure. This approach reflects a growing consensus in the field that HIV prevention and care interventions should not reproduce divisions based on HIV status. Evidence demonstrates that status-neutral programs can have better HIV and psycho-social outcomes(42). This approach both reduces stigma directly, i.e., does not reduce people to only their HIV status, and also allows for conversation among people with various experiences and perspectives to learn from each other. The curriculum integrates HIV-related self-care as part of holistic wellness but does not segregate or disproportionately emphasize HIV at the expense of other wellbeing domains. Participants responded positively to this framing, contrasting it with programs they perceived as less attractive in that they narrowly focus on risk or biomedical outcomes. HIV status neutrality also combats intra-community stigmas, fostering a more inclusive group process. As such, the Journeys program offers a template for future program adaptation in other domains where status or diagnosis can inadvertently undermine group support and program impact.

### Programmatic and Implementation Science Implications

This study underscores several implementation science principles critical for future adaptation efforts. First, early and sequential engagement with community and other stakeholders yields high-quality, context-aligned adaptations. Involving Black GBM, community leaders, and local health providers from formative research through curriculum production ensured relevance, minimized potential harms, and better equipped facilitators to manage difficult discussions about identity, trauma, and stigma. Using a recognized framework—applied flexibly—strengthens both process and outcomes. The ADAPT-ITT model provided structure, while flexibility (e.g., engaging experts throughout rather than in a single phase) supported decision-making as new themes emerged. Programs should foreground resilience, not only individual risk. Journeys centers strengths and community connectedness alongside coping and symptom reduction, aligning with a wellness-oriented approach to HIV and mental health. Finaly, implementation of Journeys demonstrated the feasibility and importance of maintaining confidentiality and ensuring robust referral pathways delivered through trusted facilitators and well-structured sessions.

### Directions for Future Research

Current pilot testing of the Journeys program will examine effects on psychosocial wellbeing and HIV service uptake among Black GBM in DC. If results are promising, program scale-up would be warranted. There is potential for adapting the Journeys program to virtual formats to address barriers of mobility, geography, and access, including in under-resourced settings. Related evaluations would employ hybrid implementation science designs both to understand effectiveness, as well as barriers and facilitators to successful scale-up. Adaptation of Journeys’ core model could also be tested among other marginalized groups, both within and beyond the US.

Future studies should directly measure internalized stigma and resilience and examine the effects of group-based learning on service uptake. More broadly, investment in partnerships between researchers and community-based organizations—and participatory approaches to adaptation, delivery, and evaluation—can accelerate both equity and impact in stigma reduction and HIV prevention/care science.

### Study Limitations

This study’s limitations include the reliance on purposive and relatively small samples for formative inputs, possibly limiting representativeness of findings. Quantitative assessment of program acceptability, feasibility, and appropriateness with larger samples will be needed as pilot and future large-scale trials proceed.

## Conclusion

Using a systematic and community-partnered approach, we successfully adapted an evidence-based program to promote mental health, HIV wellness and stigma reduction in Nigeria to the Washington, DC context. The resulting Journeys program is a promising step toward meeting local needs in culturally meaningful and sustainable ways. This model aligns with calls in HIV intervention science for status-neutral approaches, as well as those that address multiple stigmas and a shift in focus from deficits to strengths —and provides actionable lessons for both practice and research.

## Data Availability

Data underlying this manuscript, as well as data collection instruments, will be deposited in the Population Council's Harvard Dataverse repository upon acceptance of the manuscript. (https://dataverse.harvard.edu/dataverse/popcouncil)

## Acknowledgements

We thank Mario Gray, Jorian Rivera, and others who participated in facilitator training and final curriculum refinement and Alis Louviere and Katie Fan for study coordination and participant follow-up support.

